# Single dose of BNT162b2 mRNA vaccine against SARS-CoV-2 induces high frequency of neutralising antibody and polyfunctional T-cell responses in patients with myeloproliferative neoplasms

**DOI:** 10.1101/2021.04.27.21256096

**Authors:** Patrick Harrington, Hugues de Lavallade, Katie J. Doores, Amy O’Reilly, Jeffrey Seow, Carl Graham, Thomas Lechmere, Deepti Radia, Richard Dillon, Yogita Shanmugharaj, Andreas Espehana, Claire Woodley, Jamie Saunders, Natalia Curto-Garcia, Jennifer O’Sullivan, Kavita Raj, Shahram Kordasti, Michael H. Malim, Claire Harrison, Donal McLornan

## Abstract

Encouraging results have been observed from initial studies evaluating vaccines targeting the novel beta coronavirus which causes severe acute respiratory syndrome coronavirus 2 (SARS-CoV-2). However, concerns have been raised around the efficacy of these vaccines in immunosuppressed populations, including patients with haematological malignancy. Myeloproliferative neoplasms (MPN), in particular myelofibrosis (MF), are associated with heterogenous immune defects which are influenced by patient age, disease subtype and the use of cytoreductive therapies. Patients with a WHO defined diagnosis of an MPN presenting to our clinic were recruited following first injection of 30μg BNT162b2. A positive anti-S IgG ELISA was seen in 76.1% (16) of patients following vaccination with positive neutralising antibodies detected in 85.7% (18) of patients. A memory T cell response was observed in 80% (16) of patients, with a CD4+ T cell response in 75% (15) and a CD8+ T cell response in 35% (7). These results, for the first time, provide some reassurance regarding the initial immune response to the BNT162b2 vaccine amongst patients with MPN, with response rates similar to that observed in the general population.

---

Encouraging results have been observed from initial studies evaluating vaccines targeting the novel beta coronavirus which causes severe acute respiratory syndrome coronavirus 2 (SARS-CoV-2)^1,2^. BNT162b2 (Pfizer-BioNTech) is a nucleoside-modified mRNA that encodes a full-length SARS-CoV-2 Spike (S) protein, a key target of neutralising antibodies, and has demonstrated a 95% reduction of cases in the general population^1^. However, concerns have been raised around the efficacy of these vaccines in immunosuppressed populations, including patients with haematological malignancy^3^.

Myeloproliferative neoplasms (MPN), in particular myelofibrosis (MF), are associated with a pro-inflammatory state and dysregulation of pivotal natural killer cell, regulatory T cell and effector T cell function^4,5^. These heterogenous defects are further influenced by patient age, disease subtype, stage and the use of cytoreductive therapies, including JAK inhibitors^4^. A recently reported large scale population-based cohort study found incidence of both bacterial and viral infections to be significantly increased in MPN patients, irrespective of the use of cytoreductive therapies^6^. Another large patient reported prospective study evaluating incidence of infection in MPN patients found both a diagnosis of MF and the use of ruxolitinib therapy to be associated with increased risk of infection^7^. Separately, a study evaluating MF patients treated with ruxolitinib found disease severity, as determined by high international prognostic score system category, to be significantly correlated with infectious risk, with an optimal spleen response to treatment associated with improved infection free survival^8^. These studies highlight the importance of an effective vaccination programme against SARS-CoV-2 in this population. Herein we describe, for the first time, immune responses to the first injection of BNT162b2 in an unselected MPN cohort.

Patients with a WHO defined diagnosis of an MPN presenting to our clinic were recruited in accordance with the regional research and ethics review board, with sampling at baseline and median of 21 days (IQR 21-21) following first injection of 30μg BNT162b2. Clinical characteristics and adverse events are summarised in Table 1, with all adverse events reported within 7 days after administration of the vaccine considered to be related to the vaccine. The vaccine was safe and generally well tolerated with 57.1% (12) patients reporting localised inflammation and 47.6% (10) of patients reporting systemic side effects including flu-like illness, fatigue and gastrointestinal symptoms, following injection

| Pt | Age | Sex | Diagnosis | Treatment | Local side effects | Systemic side effects | Pre Anti-N IgG OD | Pre Anti-S IgG EC50 | Post Anti-S IgG EC50 | Post Neut. Ab ID50 | Post CD4+ | Post CD8+ |
| --- | --- | --- | --- | --- | --- | --- | --- | --- | --- | --- | --- | --- |
| 01 | 55-59 | F | ET | Peg-IFN | None | None | 0.317 | <25 | 25 | 95 | + | + |
| 02 | 60-64 | M | ET | Peg-IFN | None | Fatigue | 0.204 | <25 | 31 | 243 | + | + |
| 03 | 45-49 | M | MF | Surveillance | Localised inflammation | None | 0.228 | <25 | <25 | 133 | NA | NA |
| 04 | 50-54 | F | PV | Surveillance/<br>Venesection | None | None | 0.243 | <25 | <25 | <25 | + | + |
| 05 | 60-64 | F | ET | Surveillance | Localised inflammation | None | 0.207 | <25 | 25 | 156 | - | - |
| 06 | 35-39 | M | PV | HC | None | Headache,<br>Fatigue | 0.339 | <25 | 810 | 2163 | + | - |
| 07 | 40-44 | F | MF | Ruxolitinib | None | Dizziness | 0.256 | <25 | <25 | <25 | + | - |
| 08 | 60-64 | F | MF | Ruxolitinib | Localised inflammation | Fatigue | 0.900 | 25 | 5788 | 3999 | + | - |
| 09 | 40-44 | F | ET | Surveillance | None | Diarrhoea | 0.326 | <25 | 457 | 689 | - | - |
| 10 | 70-74 | M | MF | Surveillance | Localised inflammation | None | 1.772 | 41 | 9618 | 13709 | + | + |
| 11 | 60-64 | F | MF | Surveillance | Localised inflammation | None | 0.266 | 25 | 8783 | 15417 | + | + |
| 12 | 60-64 | F | ET | HC | Localised inflammation | Fatigue | 0.194 | <25 | 25 | 127 | - | - |
| 13 | 35-39 | M | MF | Ruxolitinib<br>IMG-7289 | None | None | 0.187 | <25 | 25 | 381 | + | + |
| 14 | 55-59 | F | MF | Ruxolitinib | None | None | 0.216 | <25 | <25 | <25 | + | - |
| 15 | 70-74 | F | PV | Ruxolitinib | Localised inflammation | Fatigue | 0.194 | <25 | 25 | 98 | - | - |
| 16 | 55-59 | M | ET | Peg-IFN | Localised inflammation | Flu-like illness | 0.242 | <25 | 358 | 1151 | + | - |
| 17 | 55-59 | M | MF | Ruxolitinib | None | Flu-like illness | 1.504 | 25 | 6460 | 23823 | + | - |
| 18 | 55-59 | F | ET | HC | Localised inflammation | Nausea | 0.198 | <25 | 120 | 533 | + | - |
| 19 | 40-44 | F | ET | Peg-IFN | Localised inflammation | None | 0.370 | <25 | 25 | 366 | + | + |
| 20 | 60-64 | F | MF | Hydroxy-carbamide | Localised inflammation | None | 1.739 | <25 | 457 | 1977 | + | - |
| 21 | 55-59 | F | PV | Surveillance/Venesection | Localised inflammation | None | 0.099 | NA | <25 | 213 | - | - |

EC50, 50% effective concentration; ET, essential thrombocythaemia; HC, hydroxycarbamide; ID50, 50% inhibitory dilution; MF, myelofibrosis; Peg-IFN, pegylated interferon; PV, polycythaemia vera

Anti-S IgG ELISA testing was performed as described previously^9^ in all 21 patients and results were compared with samples taken prior to vaccination in 20 patients. Neutralising antibody analysis was also performed in post-vaccine samples from all 21 patients. Briefly, HIV-1 (human immunodeficiency virus type-1) based virus particles, pseudotyped with SARS-CoV-2 Wuhan Spike were prepared in HEK-293T/17 cells and neutralization assays were conducted as previously described^10^. Serial dilutions of heat inactivated plasma samples were prepared in DMEM complete media and incubated with pseudotyped virus for 1□h at 37□°C in 96-well plates. Next, HeLa cells stably expressing the ACE2 receptor (provided by Dr James Voss, Scripps Research, La Jolla, CA) were added and the plates were left for 72 hours. Infection level was assessed in lysed cells with the Bright-Glo luciferase kit (Promega), using a Victor^™^ X3 multilabel reader (Perkin Elmer). Measurements were performed in duplicate and the duplicates used to calculate the serum dilution that inhibits 50% infection (ID_50_) using GraphPad Prism.

At baseline, 4 patients showed evidence of prior infection with positive anti-nucleocapsid IgG ELISA and an additional patient was positive for anti-S IgG. A positive anti-S IgG ELISA was seen in 76.1% (16) of patients following vaccination. The median anti-S IgG EC50 amongst positive samples was 239 (IQR 25-4544). Positive neutralising antibodies were detected in 85.7% (18) of patients, with a median ID50 of 457 (IQR 150.3-2622). Moreover, high (>501) neutralising titres were observed in 42.9% (9) of patients.

The induction of virus-specific T-cell responses by BNT162b2 vaccination was assessed *ex-vivo* by flow cytometric enumeration of antigen-specific CD8+ and CD4+ T lymphocytes using an intracellular cytokine assay for IFNγ, TNFα and IL2, as described^11^. Briefly, cells were thawed, then rested for 18 hours at 37°C, 5% CO2. Specific peptides covering the immunogenic domains of the Spike (S) protein (Miltenyi Biotech) (0.25□µg/ml) and anti-CD28 (BD bioscience) were added for 3 hours, followed by Brefeldin-A (BFA) for an additional 3 hours. Unstimulated cells were utilised as negative controls and PMA and Ionomycin (Miltenyi Biotech) was added separately as a positive control. Cells were stained with a viability dye, stained with antibodies directed against surface markers, and fixed and permeabilised (BD CytoFix/Cytoperm) prior to staining with antibodies directed against intracellular cytokines.

T cell analysis was performed in 20 patients with a response considered positive if there was a 3-fold increase in any pro-inflammatory cytokine from baseline expression, and above a threshold of 0.01. A memory T cell response was observed in 80% (16) of patients, with a CD4+ T cell response in 75% (15) and a CD8+ T cell response in 35% (7). A polyfunctional T cell response was observed in 65% (13) of patients evaluated (Figure 1 a, b,). The median increase in expression of TNFα in CD4+ cells compared with the baseline unstimulated control was 0.07 (IQR 0.01-0.35) and in CD8+ cells 0.11 (0.00-0.19). Median increase in IFNγ expression was 0.04 (−0.01-0.1) in CD4+ and 0.09 (−0.01-0.3) in CD8+ cells, whilst IL-2 was 0.05 (0.01-0.34) in CD4 and 0.02 (0.00-0.19).

**Figure 1.**
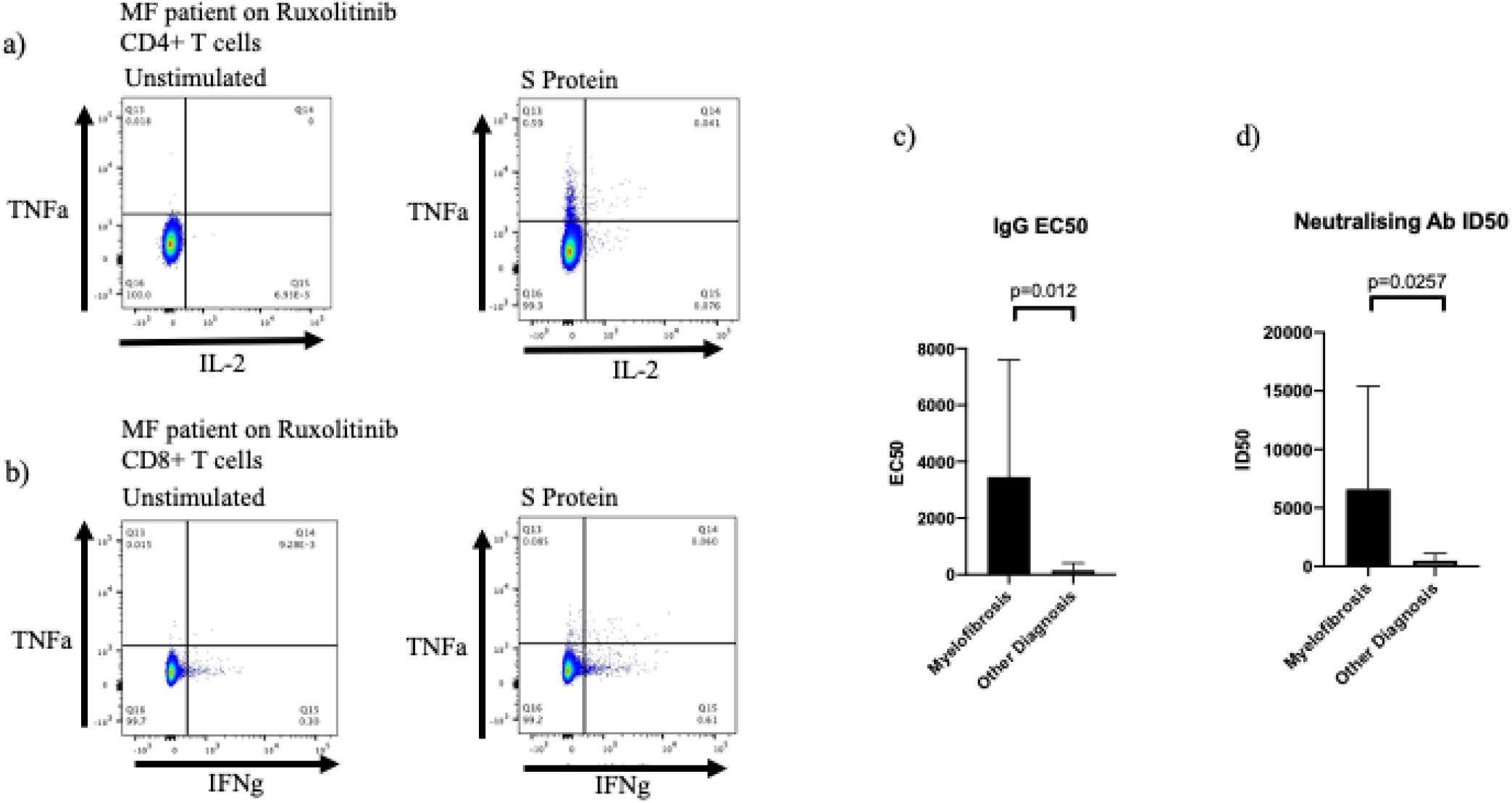
a. Post vaccine polyfunctional CD4+ T cell response in MF patient on ruxolitinib showing TNFα and IL-2 expression in unstimulated cells (left) and cells exposed to S protein (right). b. Post vaccine polyfunctional CD8+ T cell response in MF patient on ruxolitinib showing TNFα and IFNγ expression in unstimulated cells (left) and cells exposed to S protein (right). c. IgG EC50 in MF patients compared with other diagnoses. d. Neutralising antibody ID50 in MF patients compared with other diagnoses.

Of note, patients with a diagnosis of MF (n=9) had significantly higher post-vaccine anti-S IgG EC50 and neutralising antibody ID50 titres compared to patients with other MPN subtypes, with a mean IgG EC50 of 3459 vs 158.4 (p=0.012) and mean ID50 of 6604 vs 486.2 (p=0.026) respectively (Figure 1 c, d). However, 4 of the patients with evidence of previous Covid-19 infection also had a diagnosis of MF. No significant differences in T cell or antibody response were identified between patients on treatment compared with those undergoing active surveillance. Similarly, no significant differences were observed between those taking ruxolitinib, compared with other therapies.

These results, for the first time, provide some reassurance regarding the initial immune response to the BNT162b2 vaccine amongst patients with MPN, with response rates similar to that observed in the general population^12^. This is particularly relevant following reports of a reduced response to a first injection of BNT162b2 in a heterogeneous group of cancer patients, with predominantly solid tissue and lymphoid malignancies. A memory T cell response may prove to be particularly important with regards to ongoing immunity against SARS-CoV-2. Our group has demonstrated a marked decline in neutralising antibodies in the 3 months following infection^7^, whilst a robust T cell response remains evident at 6 months post infection^8^. Indeed, evidence from the SARS-CoV-1 epidemic showed the memory T cell response to be significantly more durable than antibodies^13,14^.

Further analyses of the immune response to a second injection of BNT162b2, as well as the response to other vaccines against SARS-CoV-2, are clearly required. Longitudinal studies will also need to assess the durability of these responses and confirm that vaccination translates into a reduction in cases in this population.

## Data Availability

The datasets generated during and analysed during the current study are available from the corresponding author on reasonable request.

## Acknowledgements

PH designed the research, performed the research, analysed the data and wrote the manuscript. KJD, JSe, CG, TL and MM perfomed the research and reviewed the manuscript. DR, RD, CW, JSa, NCG, JOS, KR and SK assisted with patient recruitment and reviewed the manuscript. AOR, YS and AE assisted with patient recruitment, patient interviews and reviewed the manuscript. HdL, CH and DM designed the research, assisted with patient recruitment, analysed the data and wrote the manuscript.

